# Doctors, Wellness Influencers, and Probiotic Gummies: A Cross-Sectional Analysis of Gut Health Claims and Financial Conflicts on TikTok

**DOI:** 10.64898/2026.06.15.26355698

**Authors:** Kai Vu, Kyle Garcia-Rogers, Ben Li, Vinayak Swaroop, Owais Gilani, Alice M. Tang, Michael Siegel

**Affiliations:** Department of Public Health and Community Medicine, Tufts University School of Medicine, Boston, MA, USA

**Keywords:** social media, TikTok, wellness, wellness influencers, medical influencers, health misinformation, gut health, gastroenterology

## Abstract

TikTok has emerged as a major source of health information, yet concerns persist regarding the accuracy of content and influence of financial conflicts. Gut health content is particularly vulnerable to misinformation. This study examined the relationship between creator profession (“medical” versus “non-medical”) and the quality of gut health claims and the presence of financial conflicts on TikTok. We conducted a cross-sectional study of 412 TikTok creator accounts identified using the search terms “guthealth,” “gutcleansing,” and “digestion.” One video per creator was analyzed. Creator profession was categorized as medical or non-medical. Health claim quality was coded as high, moderate, or poor. Financial conflicts (Showcase, Subscription, external links) were assessed. Modified Poisson regression was used to estimate prevalence ratios (PRs) of health claim quality (high versus poor- or moderate-quality) and financial conflicts between medical and non-medical creators, and negative binomial regression was used to evaluate associations between claim quality and number of video likes. Non-medical creators were more likely than medical creators to present poor- or moderate-quality health claims (adjusted PR: 2.33; 95% CI: 1.50–3.62). Most creators (92%) exhibited at least one financial conflict, and Showcase use was greater among non-medical creators (adjusted PR: 1.57; 95% CI: 1.02–2.42). Videos containing moderate- and poor-quality health claims received three times as many likes as videos containing high-quality claims. Non-medical creators disproportionately produced lower-quality gut health content on TikTok, and misleading claims received greater engagement. These findings highlight a misalignment between information quality and visibility, emphasizing the need for interventions promoting evidence-based health communication.

## Introduction

Social media is increasingly becoming a primary source of health-related information among adolescents and young adults.^1^ TikTok, a platform with nearly 1.9 billion monthly users as of early 2026, the majority of whom are between the ages of 18 and 34, has emerged as one of the most influential apps shaping health beliefs and behaviors.^2^ Previous studies estimate that anywhere from 25% to over 75% of adolescents and young adults view social media as a useful source of health information, often supplementing or replacing guidance from health professionals.^3,4^ While social media platforms offer new opportunities for public health communication, growing evidence suggests that much of the health-related content circulated online is not evidence-based and is frequently disseminated by non-credentialed individuals.^5–8^ Nutrition- and digestive-health-related content has been identified as a particularly high-risk domain for misinformation, with viral videos commonly promoting detoxification practices, supplements, and restrictive diets that lack robust clinical evidence.^9^ Exposure to misleading digestive health content may influence dietary behaviors, supplement use, and healthcare-seeking patterns, with potential downstream clinical consequences.

Previous analyses of TikTok health content demonstrate that misleading or unverifiable claims made by creators often achieve equal or greater engagement than evidence-based information, presenting a major public health concern.^10,11^ These patterns are not specific to creators within the general public, as physicians and other medical creators have been shown to receive greater audience engagement from content containing poorer-quality health claims.^11^ Many social media algorithms are designed to maximize engagement, which systematically privileges content that triggers attention – often moral/emotional and divisive content that overlaps with misinformation.^12^ Popularity- and relevance-based platform algorithms have been identified as major drivers of online misinformation dissemination.^13,14^

Despite growing recognition of this issue, important gaps remain in the literature. A limited number of papers have evaluated the quality of gastrointestinal health content on social media platforms.^15–18^ Past studies have relied on small samples using a single search term, with limited evaluation of creator demographics, professional background, and various markers of professional credibility.^5^ Furthermore, little research has examined how monetization strategies like commission-based affiliate marketing, external product promotion, or subscriptions vary across creator professions (e.g., “medical” versus “non-medical” creators) on TikTok, nor how this may influence the framing and credibility of health claims. Together, these limitations restrict understanding of how creator identity and financial conflicts interact to shape the quality and dissemination of health information on TikTok.

To address these gaps, we conducted a cross-sectional study of TikTok videos related to gut health using multiple search queries and a standardized manual coding protocol. We examined whether creator professional background (medical versus non-medical) was associated with the quality of health claims presented and with the presence of financial conflicts on creator profiles. Additionally, we compared creator characteristics and platform features across professional groups to better understand differences in audience reach and presentation strategies. Lastly, we assessed whether the quality of health claims in videos was associated with the number of likes received. We hypothesized that non-medical creators would be more likely to present lower-quality health claims and to utilize monetization features compared with medical creators, and that lower-quality health claims would attract greater audience engagement.

## Methods

### Data Collection and Study Eligibility

Data collection followed a standardized process. First, a new TikTok account was created with no prior search or watch history to minimize personalization of search results and reduce potential algorithmic bias from prior user activity. Creator usernames were then identified through platform search queries using the terms “guthealth,” “gutcleansing,” and “digestion.” Searches were conducted on December 12, 2025 (“guthealth,” “gutcleansing”) and January 20, 2026 (“digestion”). This third search query was conducted to increase representation of medical creators identified through platform searches. Usernames returned from each search were copied into separate lookup sheets stratified by search term. Duplicate accounts appearing within or across search queries were identified and removed prior to data extraction. Each unique username was assigned a numeric study ID, and the username-ID linkage file was stored separately from the analytical dataset.

One of four independent reviewers accessed each account using the TikTok mobile application and manually recorded observational data from the indexed video and corresponding creator profile in a dataset containing only study IDs and study variables. To assess interrater reliability, a randomly selected 10% subsample of accounts was independently reviewed by all four reviewers. Data collection began on December 20, 2025, and was completed on February 16, 2026.

Inclusion criteria were: (1) publicly accessible TikTok creator accounts identified through predefined search queries, (2) accounts representing individual content creators, and (3) accounts with at least 5,000 followers. Accounts for which the creator or indexed video could not be located, as well as those with unavailable or unrelated profession information, were subsequently excluded.

### Measures

Data were collected on a combination of video- and account-level variables for each creator. For each video identified in our search, the number of likes, date posted, and whether or not subtitles were present were recorded. Video dates were further categorized by calendar year and quarter (i.e., January to March, April to June, etc.). The following creator account-level characteristics associated with each respective video were recorded: observed gender (man, woman, indeterminate/other), observed age (< 40 or ≥ 40), verification status (whether or not the user had an official TikTok verification badge), follower count, total like count across all account videos, and whether or not the full name and/or “Doctor” (or “Dr.”) prefix was used in the account name, username, or biography.

Creator profession was coded based on the credentials, title, and role presented on the creator’s TikTok profile, username, biography, linked content, and other accessible public-facing materials. Profession categories included physicians, nurse practitioners, scientists (including researchers and professors), dentists, other professionally prepared clinicians, chiropractors, alternative and functional medicine specialists, and health coaches and influencers. Physicians were defined as both US (MD and DO) and international (MD, MBBS, MBChB, etc.) medical graduates. “Other professionally prepared clinicians” were defined as clinical professions requiring professional or graduate degrees, and included physician assistants, occupational therapists, physical therapists, podiatrists, optometrists, perfusionists, licensed therapists, mental health counselors, social workers, audiologists, and registered dietitians. “Alternative or functional medicine specialists” included naturopathic doctors, naturopaths, functional medicine specialists, acupuncturists, and related alternative providers. “Health coaches and influencers” included wellness influencers, coaches, personal trainers, and nutritionists who were not registered dietitians. Professions were recoded dichotomously for analysis as medical creators (physicians, nurse practitioners, dentists, and other professionally prepared clinicians) or non-medical creators (chiropractors, alternative and functional medicine specialists, and health coaches and influencers). Licensure or certification was determined by whether the creator presented evidence of professional licensure, board certification, or other professional credentials on their profile.

Each video was reviewed to assess the quality of the health claim present and was coded in one of four categories: poor, moderate, high, or no claim present. Poor-quality claims were characterized by harmful or misleading information and were defined as unsubstantiated or overstated medical claims lacking supporting evidence, contradicting accepted guidelines, relying on anecdotes or pseudoscience, using fear-mongering, or lacking biological plausibility. Moderate-quality claims were defined as partially substantiated or plausible claims that were exaggerated or overgeneralized based on weak evidence or lacking appropriate context regarding risks or limitations. High-quality claims were defined as well-substantiated and evidence-based claims presented responsibly with acknowledgment of limitations when applicable and without exaggeration. Health claim quality was further recoded into a dichotomous variable (high- versus low- or moderate-quality) for some analyses. In cases of reviewer uncertainty regarding health claim quality, OpenEvidence was used to evaluate the claim against current literature, inputting the exact wording of the creator in the OpenEvidence search query.^19^ Videos with content unrelated to health or that had no health-relevant takeaways were categorized as no claim present.

Creator financial conflicts were assessed using three separate dichotomous measures (present or not present) directly from the account profile page: “Showcase,” “Subscription,” and the presence of external links with potential for monetization. Showcase was coded as present if the creator had a TikTok Showcase available on their profile, which allows creators to receive commission from linked product purchases (either directly from videos or from the account profile) through TikTok Shop. Subscription was coded as present if the creator had a Subscription available on their profile, an optional paid feature that viewers can purchase to receive access to exclusive videos and tiered badges, among other perks from the creator. External monetization was coded as present if the creator included links to external websites, including but not limited to Linktree, Shopify, merchandise, personal websites, courses, appointments, or blog posts. These variables were also combined into a dichotomous composite variable indicating the presence of at least one monetization method.

### Statistical Analysis

All analyses were performed in R version 4.5.2.^20^ Creator account and video characteristics were summarized descriptively. Bivariate analyses compared select characteristics by creator profession (i.e., non-medical versus medical) using the Wilcoxon Rank-Sum test and chi-squared test for continuous and categorical variables, respectively. Select characteristics, including doctor prefix, verification status, full name usage, and licensure or certification, were included as indicators of professional authority and credentials. All statistical tests were two-sided, and statistical significance was defined as α = 0.05.

Our primary objective was to assess the relationship between creator profession and health claim quality. Health claim quality was analyzed dichotomously (poor- or moderate- versus high-quality). We generated crude and adjusted prevalence ratios (PRs), comparing the prevalence of poor- and moderate- versus high-quality health claims by creator profession. Estimates were calculated using Poisson regression with robust standard errors. The adjusted model controlled for age, gender, year, quarter, and search query. These variables were selected to minimize confounding from creator demographics (age and gender), temporal trends and seasonality (year and quarter), and differing distributions of claim quality varying by search query.

We also assessed the association between creator profession and financial conflicts. Crude and adjusted models were fit using four measures of financial conflicts: Showcase, Subscription, external link, and any of the aforementioned. Our adjusted models again controlled for age, gender, year, quarter, and search query.

Finally, we examined the association between health claim quality and the number of video likes received. Crude and adjusted negative-binomial regression models were fit to estimate incidence rate ratios (IRRs) for video likes across claim quality categories. In this analysis, health claim quality was modeled as a three-level categorical variable, with high-quality claims as the reference category. The adjusted model controlled for age, gender, year, quarter, search query, number of account followers, presence of subtitles in the video, and Showcase presence. The latter three variables were included due to being visible at the video level and thus potentially confounding this association. Lastly, we conducted an interaction analysis to assess whether the association between health claim quality and video likes differed between medical and non-medical creators.

Interrater reliability was assessed using Fleiss’ kappa (κ) among a 10% subsample coded individually by all four reviewers for select categorical variables requiring subjective interpretation. To account for potential misclassification bias, two post hoc sensitivity analyses were performed: one excluding unverified “doctors” from the non-medical creator subgroup and another categorizing unverified “doctors” as medical creators, with primary and secondary objective analyses repeated under both approaches.

### Ethical Approval

The study protocol was reviewed by the Tufts Health Sciences Institutional Review Board and determined not to constitute human subjects research (STUDY00006621).

## Results

### Sample Identification and Study Population

A total of 509 TikTok creator accounts were initially identified using the search queries “guthealth,” “gutcleansing,” and “digestion” (Figure 1). After duplicate removal, 412 unique creator accounts were screened for eligibility. Of these, 238 accounts were excluded because the creator or indexed video could not be identified (n = 41), the account was not an individual creator account (n = 64), profession was unavailable or unrelated (n = 100), or the creator had fewer than 5,000 followers (n = 33). The final analytic sample included 174 creators.

**Figure 1.**
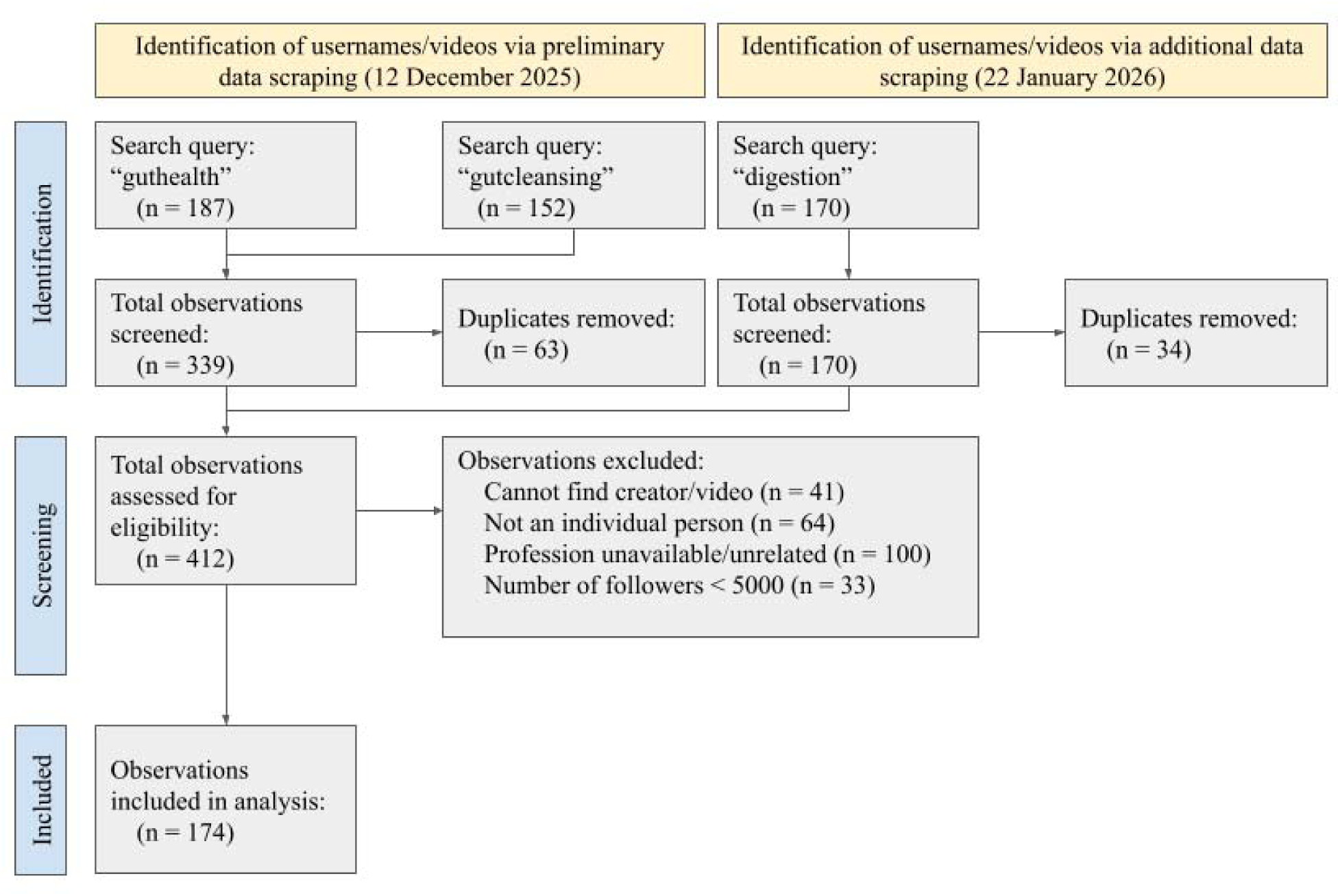
Data collection timeline and study selection flowchart.

### Creator Account and Video Characteristics

Out of 174 TikTok creators in the final sample, 29 (17%) were medical creators and 145 (83%) were non-medical creators (Table 1). Among medical creators, the most common professions were physicians (52%) and other professionally prepared clinicians (28%), whereas the overwhelming majority of non-medical creators were health coaches and influencers (90%). A detailed list of credentials by profession is provided in the Supplementary Information (Supplementary Table 1).

**Table 1.**
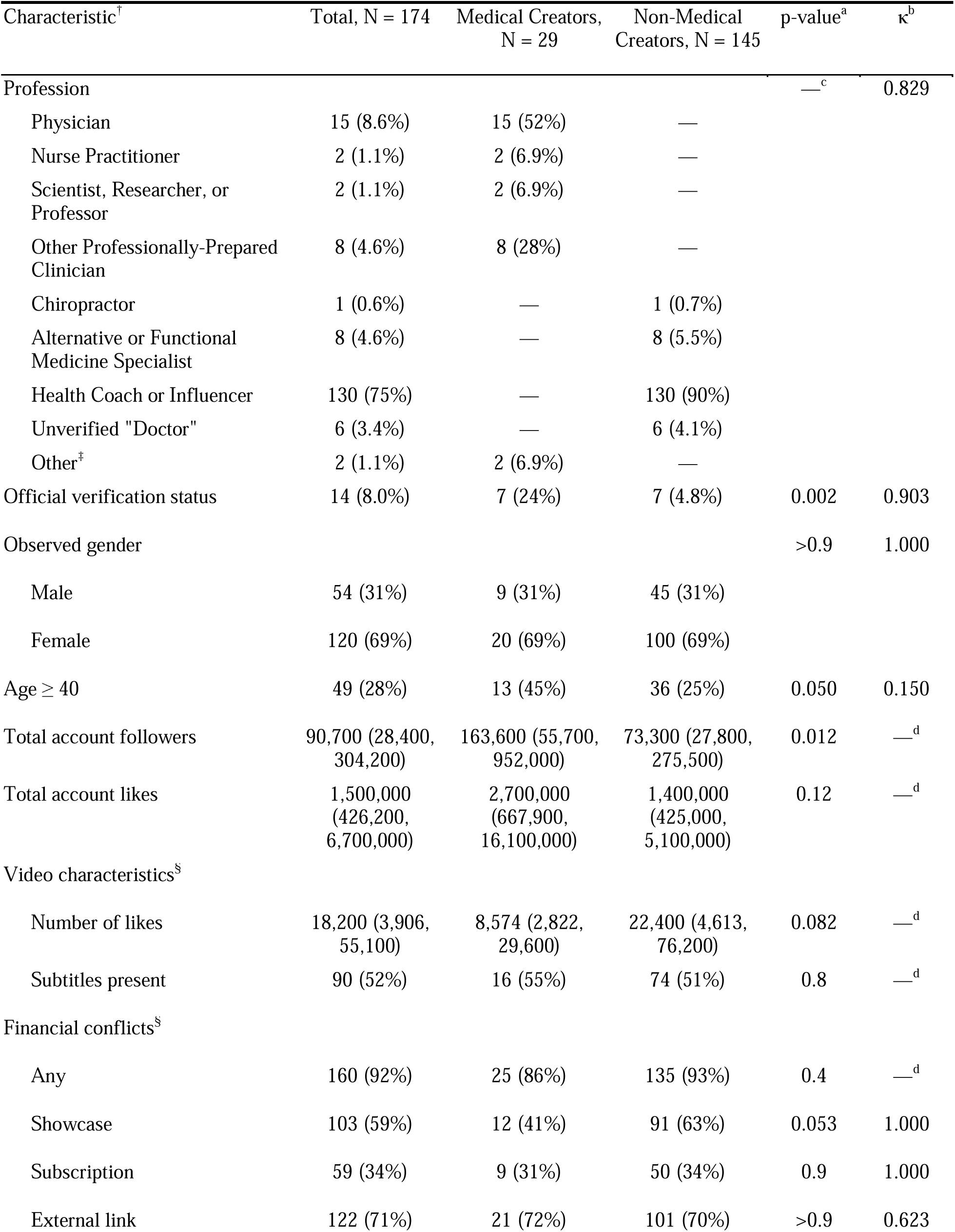
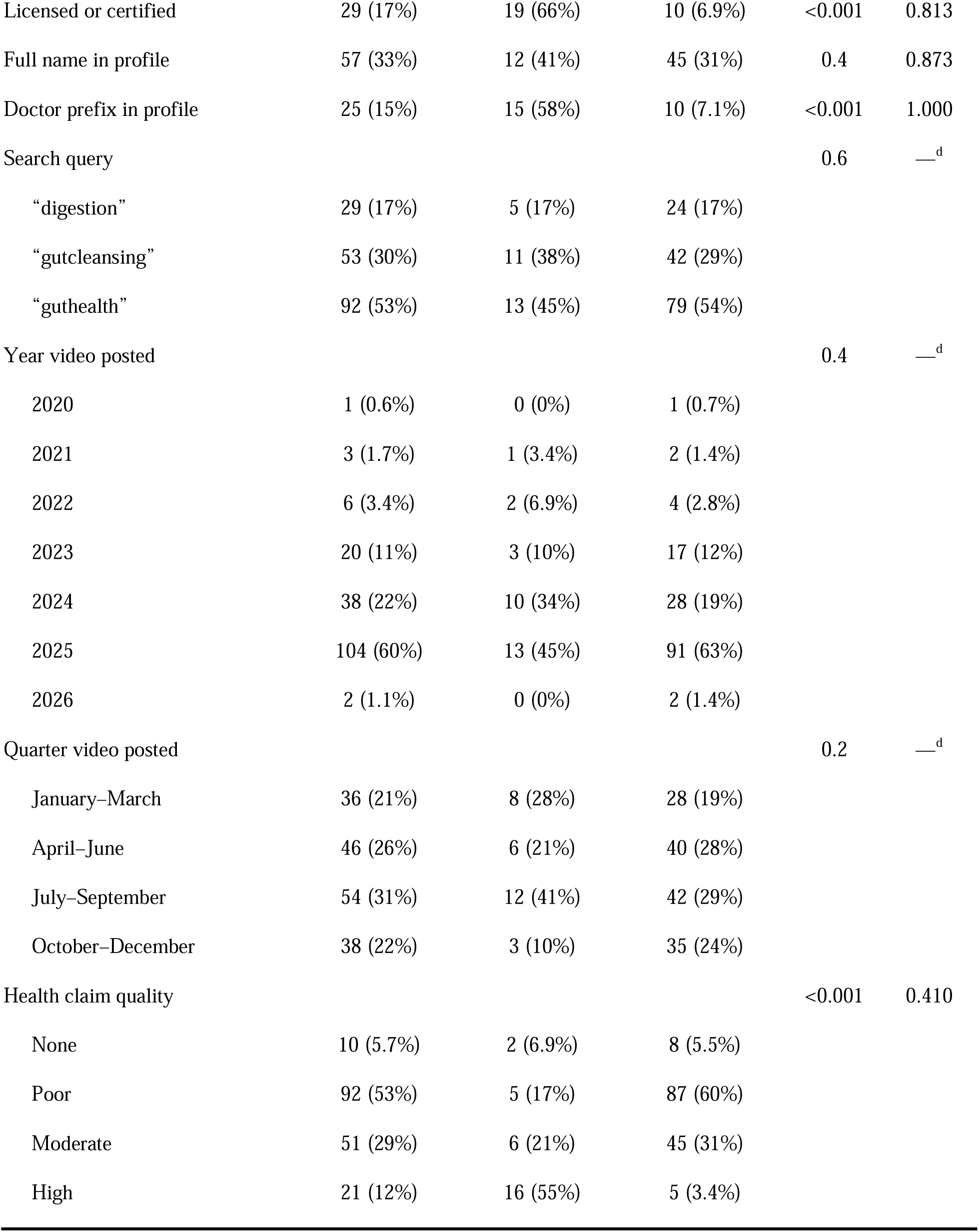
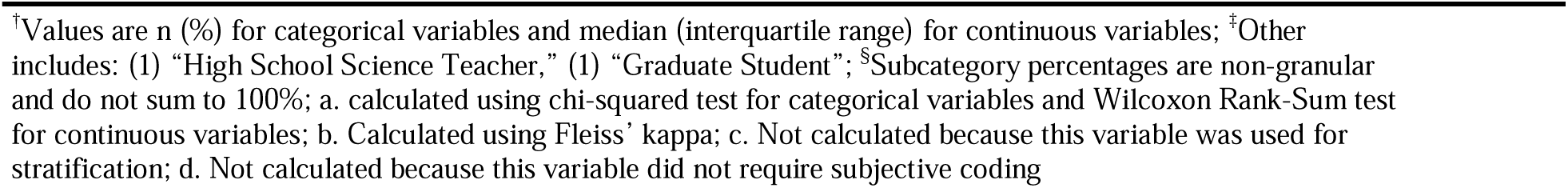
Characteristics by creator profession (N = 174)

Medical creators were more likely than non-medical creators to have verification badges, list licensure or certification, and use a doctor prefix in their profiles (24% vs 4.8%, p = 0.002; 66% vs 6.9%, p < 0.001; 58% vs 7.1%, p < 0.001). Medical creators also had a higher median follower count than non-medical creators (163,600 vs 73,300, p = 0.012). Median total account likes were higher among medical creators, but did not differ statistically significantly between groups. Financial conflicts were highly prevalent and did not differ between groups (86% vs 93%, p = 0.4).

Indexed videos differed statistically significantly in health claim quality by creator type (p < 0.001): high-quality claims were most common among medical creators (55%), whereas poor-quality claims were most common among non-medical creators (60%). Interrater reliability was high overall across subjectively coded variables, with most demonstrating moderate to almost perfect agreement across four reviewers (κ range: 0.15–1.00). Only one variable, age, demonstrated slight agreement (κ = 0.15). Agreement for health claim quality was moderate (κ = 0.41).

### Creator Profession and Health Claim Quality

Compared with medical creators, non-medical creators were more than twice as likely to produce poor- or moderate-quality health claims in their videos (Table 2; adjusted prevalence ratio [PR]: 2.33; 95% confidence interval [CI]: 1.50–3.62). Among 137 non-medical creators, only five (3.6%) produced high-quality health claims.

**Table 2.**
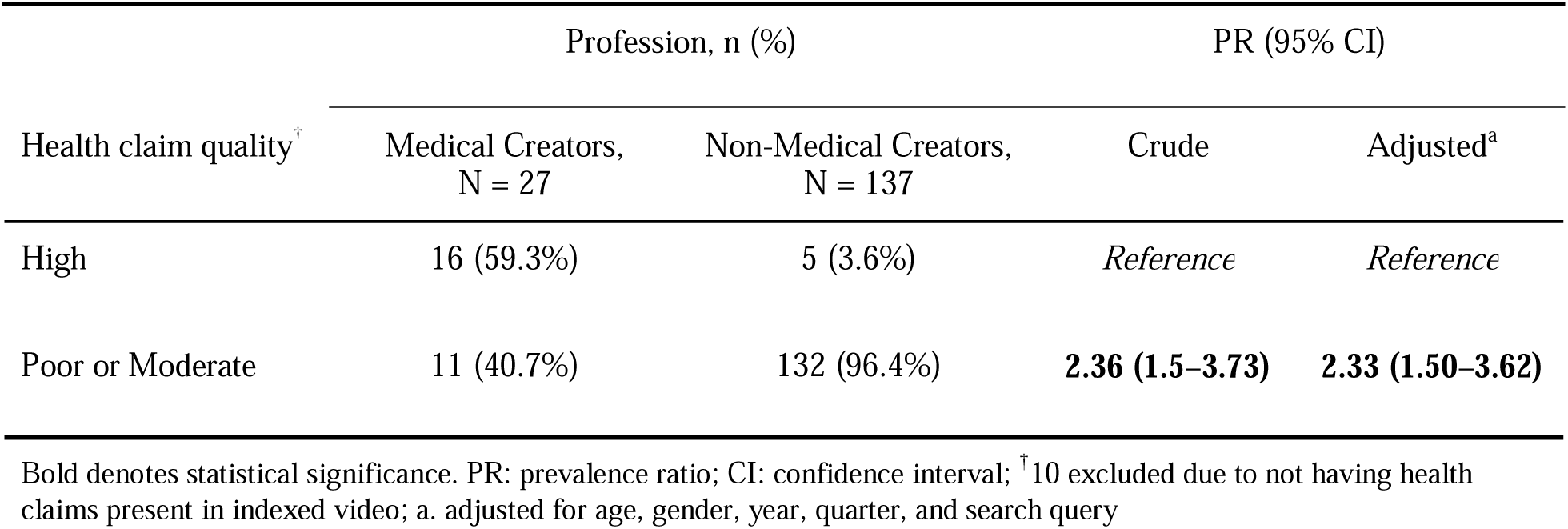
Association between creator profession and health claim quality (N = 164)

| Health claim quality<sup>†</sup> | Profession, n (%) |  | PR (95% CI) |  |
| --- | --- | --- | --- | --- |
|  | Medical Creators,<br>N = 27 | Non-Medical Creators,<br>N = 137 | Crude | Adjusted<sup>a</sup> |
| High | 16 (59.3%) | 5 (3.6%) | *Reference* | *Reference* |
| Poor or Moderate | 11 (40.7%) | 132 (96.4%) | **2.36 (1.5–3.73)** | **2.33 (1.50–3.62)** |

Bold denotes statistical significance. PR: prevalence ratio; CI: confidence interval; <sup>†</sup>10 excluded due to not having health claims present in indexed video; a. adjusted for age, gender, year, quarter, and search query

### Creator Profession and Financial Conflicts

Compared with medical creators, non-medical creators had a higher prevalence of the Showcase feature on their profiles (Table 3; adjusted PR: 1.57; 95% CI: 1.02–2.42). The adjusted association was statistically significant, whereas the crude association was not. No associations were identified between creator profession and the presence of subscriptions or external links.

**Table 3.** Adjusted associations between creator profession and financial conflicts (N = 174)

| Financial conflict | Profession, n (%) |  | PR (95% CI) |  |
| --- | --- | --- | --- | --- |
|  | Medical Creators,<br>N = 29 | Non-Medical Creators,<br>N = 145 | Crude | Adjusted<sup>a</sup> |
| Any | 25 (86.2%) | 135 (93.1%) | 1.08 (0.93–1.26) | 1.09 (0.94–1.27) |
| Showcase | 12 (41.4%) | 91 (62.8%) | 1.52 (0.97–2.38) | **1.57 (1.02–2.42)** |
| Subscription | 9 (31%) | 50 (34.5%) | 1.11 (0.62–2.00) | 1.14 (0.66–1.98) |
| External link | 21 (72.4%) | 101 (70.3%) | 0.97 (0.76–1.24) | 0.98 (0.76–1.28) |

Bold denotes statistical significance. PR: prevalence ratio; CI: confidence interval; a. adjusted for age, gender, year, quarter, and search query

### Health Claim Quality and Video Engagement

Compared with high-quality claims, videos containing moderate- or poor-quality claims received over three times as many likes (Table 4). Specifically, moderate-quality claims were associated with 3.16 times more likes (adjusted incidence rate ratio [IRR]: 3.16; 95% CI: 1.30–7.26), and poor-quality claims were associated with 3.37 times more likes (adjusted IRR: 3.37; 95% CI: 1.41–7.51). No statistically significant interaction was observed between creator profession and health claim quality in relation to video likes.

**Table 4.**
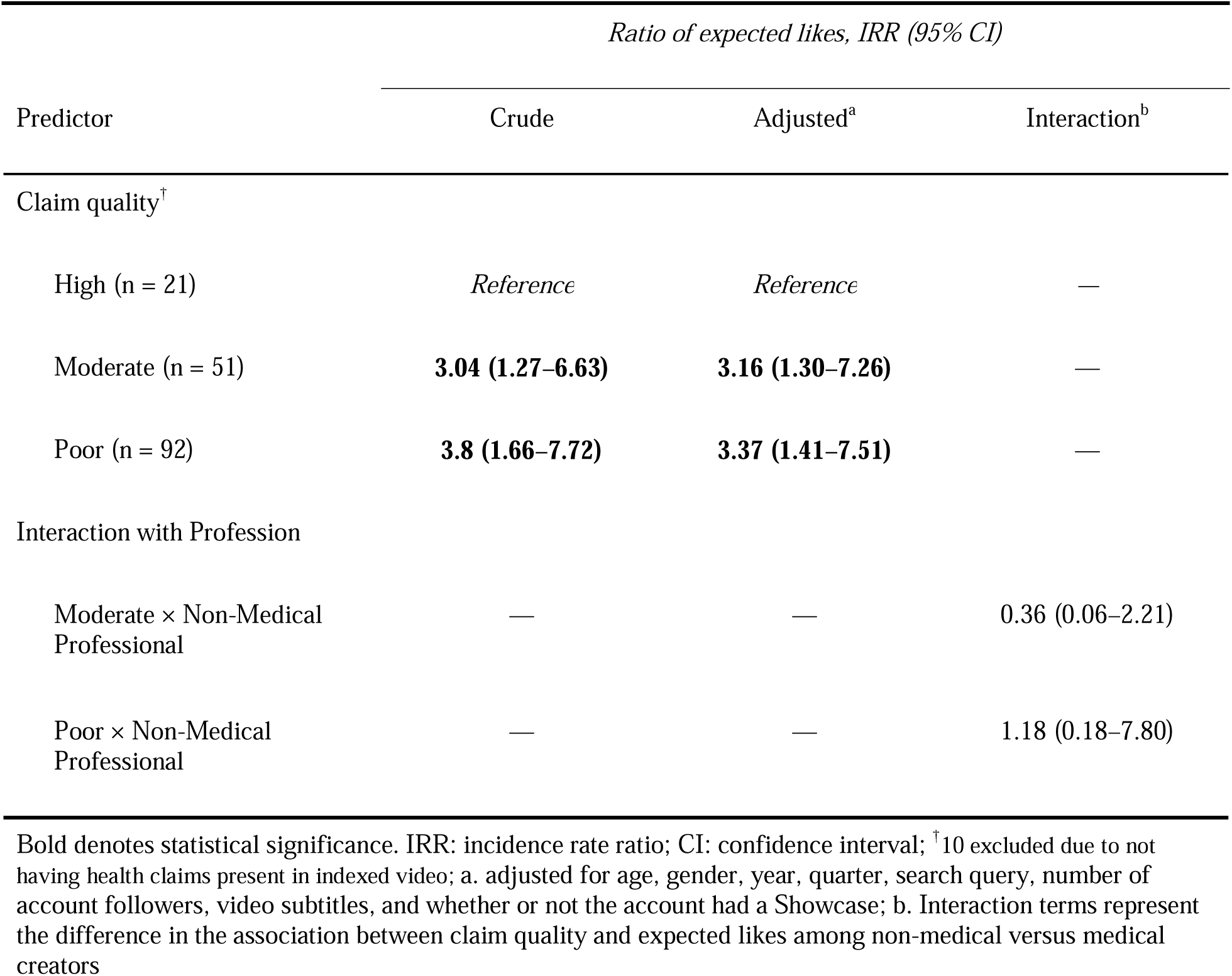
Association between health claim quality and video likes (N = 164)

| Predictor | *Ratio of expected likes, IRR (95% CI)* |  |  |
| --- | --- | --- | --- |
|  | Crude | Adjusted<sup>a</sup> | Interaction<sup>b</sup> |
| Claim quality<sup>†</sup> |  |  |  |
| High (n = 21) | *Reference* | *Reference* | — |
| Moderate (n = 51) | **3.04 (1.27–6.63)** | **3.16 (1.30–7.26)** | — |
| Poor (n = 92) | **3.8 (1.66–7.72)** | **3.37 (1.41–7.51)** | — |
| Interaction with Profession |  |  |  |
| Moderate × Non-Medical Professional | — | — | 0.36 (0.06–2.21) |
| Poor × Non-Medical Professional | — | — | 1.18 (0.18–7.80) |

Bold denotes statistical significance. IRR: incidence rate ratio; CI: confidence interval; <sup>†</sup>10 excluded due to not having health claims present in indexed video; a. adjusted for age, gender, year, quarter, search query, number of account followers, video subtitles, and whether or not the account had a Showcase; b. Interaction terms represent the difference in the association between claim quality and expected likes among non-medical versus medical creators

### Sensitivity Analysis

Sensitivity analyses excluding six unverified “doctors” yielded findings similar to the primary analyses; however, the crude association between creator profession and Showcase use became statistically significant (Supplementary Tables 2 and 3; crude PR: 1.58; 95% CI: 1.01–2.48). When unverified “doctors” were recategorized as medical creators, the magnitude of the association between creator profession and health claim quality decreased slightly (Supplementary Table 4; adjusted PR: 1.88; 95% CI: 1.37–2.58). Additionally, both the crude and adjusted associations between profession and any financial conflict present became statistically significant, with non-medical creators being more likely to have any form of analyzed financial conflict present (Supplementary Table 5; adjusted PR: 1.24; 95% CI: 1.05–

1.48).

## Discussion

Our study provides evidence that non-medical creators are more likely to present poorer-quality health claims and use commission-based affiliate marketing compared to medical creators within the gut health and wellness space on TikTok. This cross-sectional analysis is, to our knowledge, the first to directly analyze differences in health claim quality and financial conflicts by creator profession in social media content. Additionally, videos containing poorer-quality gut health claims received over three times as many likes compared to high-quality claims.

These findings are concerning given the scale of the TikTok user base and the potential for large-scale dissemination of misinformation, particularly for viewership and financial gain. Moreover, the relationships between creator profession, health claim quality, and video engagement suggest that creator, audience, and algorithmic preferences may jointly shape the visibility of health information online; videos containing poorer-quality health claims may be produced by creators who rely on TikTok for financial income and thus be more likely to utilize more effective engagement-oriented strategies. This highly engaging content may be further amplified by platform recommendation systems to maximize user engagement and platform revenue, regardless of the accuracy of the information presented.

Interestingly, we did not observe a statistically significant effect of profession on the association between health claim quality and video likes. However, this may reflect limited statistical power, as only 5 and 6 medical creators had poor- and moderate-quality health claims, respectively.

Previous studies have demonstrated this negative association between health claim quality and audience engagement among both medical and non-medical creators.^10,11^ Whether the strength of this relationship differs between groups remains unclear and warrants further investigation.

Although most creators in our sample exhibited at least one financial conflict, we observed only one statistically significant association between creator profession and monetization: non-medical creators were more likely than medical creators to list a Showcase in their profile (63% vs. 42%). External links were included as a proxy for potential monetization; however, not all links directly generate revenue. For example, a medical creator linking to the Centers for Disease Control and Prevention may be classified as having an external link despite having no financial intent. This may have resulted in misclassification and attenuated associations involving this measure. Similarly, TikTok Subscriptions were initially introduced in 2022 as an invite-only feature and only became widely available near the end of 2024.^21^ As a relatively new and evolving feature, their uptake remains limited, and associations with creator profession may be unstable and require reassessment over time. In contrast, Showcases – which allow creators to link products from TikTok Shop and earn commissions – represent a more direct measure of monetization. Given the growing integration of TikTok Shop into the platform and its role in facilitating product-based sales, Showcase may be particularly relevant in gut-health content, where supplements and wellness products are commonly marketed. While our measure captured the presence of a Showcase at the profile level rather than whether the identified video included a product link, this likely reflects a creator’s overall engagement in product-based monetization. Taken together, these considerations suggest that Showcases may represent a more robust indicator of financial conflicts, supporting the observed association with creator profession.

This study has several limitations. Health claim quality was assessed using a structured coding framework developed for this study rather than a previously validated tool. Few validated instruments have been used to evaluate short-form health content; although tools such as DISCERN have been utilized in other studies analyzing short-form content, these tools were designed and validated specifically for written health information.^22^ To our knowledge, no validated scale currently exists specifically for evaluating health claims in short-form video. Although this introduces potential misclassification bias in health claim quality assessment, interrater reliability for health claim quality was moderate (κ = 0.41) and comparable to the original DISCERN validation study, which reported weighted κ values of 0.53 among expert panelists and 0.40 among professional information providers.^23^ Despite differences in κ statistics, both measure chance-corrected agreement, supporting the internal consistency of our approach while highlighting the inherent subjectivity of evaluating health information quality.

Reviewer bias may have influenced the classification of health claims, with reviewers potentially appraising content from medical creators more favorably. Therefore, our coding approach emphasized biological plausibility, support from peer-reviewed literature, and potential for harm rather than strict concordance with clinical guidelines alone. Moreover, health claim quality was assessed before collecting account-level characteristics, limiting potential bias to videos in which creators explicitly disclosed their profession. Selection bias is also a concern given our sampling strategy. Duplicate creators appearing within and across search queries were removed so that each creator contributed only one video to the analytic sample. This approach prevented highly visible creators from being overrepresented, but only the first indexed video from each creator was analyzed. As a result, the selected video may not reflect the overall quality or tone of the creator’s broader content.

In addition, the sample was limited to publicly accessible individual creator accounts with at least 5,000 followers identified through predefined search terms. Different gut-health-related search queries, smaller creators, private accounts, or organizational accounts may have produced different distributions of creator professions, health claim quality, financial conflicts, and engagement. Furthermore, unverified “doctors” – creators using “Doctor” or “Dr.” prefixes without easily identifiable evidence of medical training – were categorized as non-medical, potentially introducing misclassification bias. To address this concern, we conducted sensitivity analyses excluding these creators, which did not reveal appreciable differences in our results. However, residual misclassification remains possible when professional identity could not be fully confirmed from public information. Additionally, measures of financial conflicts were also limited to visible, profile-level features and may not reflect undisclosed sponsorships or other revenue streams outside TikTok, thus potentially underestimating creators’ financial conflicts.

TikTok’s search and recommendation systems are proprietary, dynamic, and not fully reproducible.^24^ We attempted to mitigate the “black-box” nature of the algorithm by minimizing personalization; new accounts were created with no prior search or watch history. However, search results may still have been influenced by geographical factors, device-level signals, timing, or other algorithmic features. Because TikTok accounts require an email address for registration, the use of a preexisting email account with an institution-affiliated domain could have potentially introduced residual personalization or algorithmic bias. Accordingly, the analyzed content may not be representative across geographic, temporal, or algorithmic contexts. Content generalizability is also limited because gut health and wellness represent a particularly monetizable niche in which supplements, cleansing, detoxification, and microbiome-related claims are common. The effect size of our associations may not generalize to other lucrative areas of digital health misinformation, such as dermatology, psychiatry, or reproductive health; further research is needed to evaluate the quality of online health information in these areas. Similarly, results may not generalize to other platforms, including Instagram, YouTube, or long-form websites, where content length, user engagement, and monetization potential differ substantially.

Given research estimating that over three-quarters of youth rely on social media as a trusted source of health information, more resources should be allocated toward surveillance and interventions to mitigate online health misinformation.^4^ This could include automated fact-checking systems for videos flagged as potentially containing health information, with explicit labeling of videos when misinformation is present. Social media platforms like TikTok may also benefit from additional verification badges or labels for licensed medical professionals. Platforms may further promote evidence-based content through dedicated health-information sections featuring verified medical professionals, as has been implemented elsewhere.^25^

## Conclusion

Non-medical creators, who comprised the majority of gut health content on TikTok, were significantly more likely to present poor- or moderate-quality health claims compared with medical creators. Despite this, lower-quality content was associated with greater engagement, suggesting that platform dynamics may preferentially amplify misleading information. Financial conflicts were highly prevalent across creator types, with commission-based affiliate marketing being more commonly used among non-medical creators. Together, these findings highlight a misalignment between the quality and visibility of health information on social media platforms. Interventions such as automated fact-checking, improved credential verification, and algorithmic prioritization of evidence-based content may help mitigate the spread of misinformation and promote higher-quality online health information.

## List of abbreviations

CI: Confidence interval
IRR: Incidence rate ratio
PR: Prevalence ratio
κ: Kappa statistic

## Declarations

### Consent for publication

Not applicable.

### Data Availability

The data that support the findings of this study are openly available in Harvard Dataverse at https://doi.org/10.7910/DVN/IYNN0A.

### Authors’ Contributions

KV conceptualized the study, developed the study design, led data collection and analysis, and drafted the manuscript. KGR contributed to the study design, participated in data collection and analysis, and co-drafted the manuscript. BL and VS contributed to the study design, participated in data collection and analysis, and assisted in writing the manuscript. OG and AT provided statistical and methodological guidance and contributed to manuscript revision. MS supervised the study and provided senior oversight and critical revision of the manuscript. All authors reviewed and approved the final manuscript prior to submission.

## Acknowledgements

Not applicable.

---

**Supplementary Table 1.** Credentials by creator profession (N = 174)

| Profession<sup>†</sup> | Total,<br>N = 174 | Medical Creators,<br>N = 29 | Non-Medical Creators,<br>N = 145 |
| --- | --- | --- | --- |
| Physician | 5 (2.9%) | 5 (17%) | — |
| Physician (DO) | 2 (1.1%) | 2 (6.9%) | — |
| Physician (MD) | 7 (4.0%) | 7 (24%) | — |
| Physician (MD, FACS) | 1 (0.6%) | 1 (3.4%) | — |
| Scientist/Researcher/Professor | 1 (0.6%) | 1 (3.4%) | — |
| Scientist/Researcher/Professor (PhD, BSc, BEd) | 1 (0.6%) | 1 (3.4%) | — |
| Nurse Practitioner | 1 (0.6%) | 1 (3.4%) | — |
| Nurse Practitioner (FNP) | 1 (0.6%) | 1 (3.4%) | — |
| Other Professionally-Prepared Clinician | 6 (3.4%) | 6 (21%) | — |
| Other Professionally-Prepared Clinician (PT, BS) | 1 (0.6%) | 1 (3.4%) | — |
| Other Professionally-Prepared Clinician (RDN) | 1 (0.6%) | 1 (3.4%) | — |
| Other<sup>‡</sup> | 2 (1.1%) | 2 (6.9%) | — |
| Health Coach/Influencer | 126 (72%) | — | 126 (87%) |
| Health Coach/Influencer (BS) | 1 (0.6%) | — | 1 (0.7%) |
| Health Coach/Influencer (LMT) | 1 (0.6%) | — | 1 (0.7%) |
| Health Coach/Influencer (MOMsSc) | 1 (0.6%) | — | 1 (0.7%) |
| Health Coach/Influencer (TTS) | 1 (0.6%) | — | 1 (0.7%) |
| Alternative/Functional Medicine Specialist | 7 (4.0%) | — | 7 (4.8%) |
| Alternative/Functional Medicine Specialist (ND) | 1 (0.6%) | — | 1 (0.7%) |
| Chiropractor (DC) | 1 (0.6%) | — | 1 (0.7%) |
| Unverified "Doctor" | 5 (2.9%) | — | 5 (3.4%) |
| Unverified "Doctor" (MD) | 1 (0.6%) | — | 1 (0.7%) |

<sup>†</sup>Values are n (%); <sup>‡</sup>Other includes: (1) "High School Science Teacher," (1) "Graduate Student"

**Supplementary Table 2.**
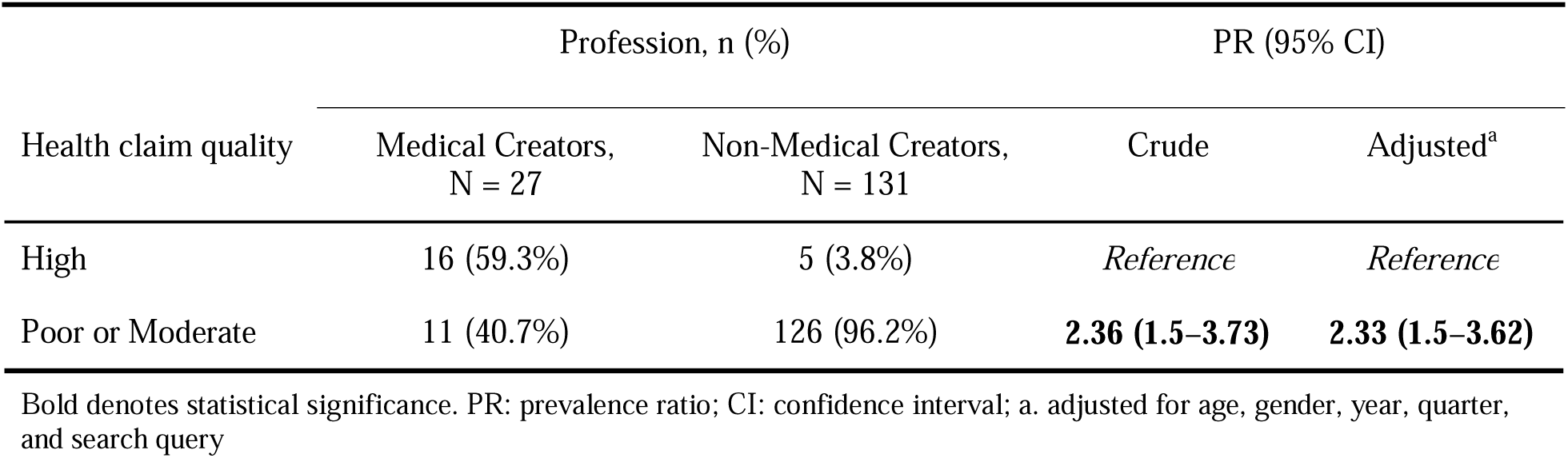
Association between creator profession and health claim quality without unverified “doctors” (N = 158)

| Health claim quality | Profession, n (%) |  | PR (95% CI) |  |
| --- | --- | --- | --- | --- |
|  | Medical Creators,<br>N = 27 | Non-Medical Creators,<br>N = 131 | Crude | Adjusted<sup>a</sup> |
| High | 16 (59.3%) | 5 (3.8%) | *Reference* | *Reference* |
| Poor or Moderate | 11 (40.7%) | 126 (96.2%) | **2.36 (1.5–3.73)** | **2.33 (1.5–3.62)** |

Bold denotes statistical significance. PR: prevalence ratio; CI: confidence interval; a. adjusted for age, gender, year, quarter, and search query

**Supplementary Table 3.** Associations between creator profession and financial conflicts without unverified “doctors” (N = 168)

| Financial conflict | Profession, n (%) |  | PR (95% CI)<sup>a</sup> |  |
| --- | --- | --- | --- | --- |
|  | Medical Creators,<br>N = 29 | Non-Medical Creators,<br>N = 139 | Crude | Adjusted<sup>a</sup> |
| Any | 25 (86.2%) | 133 (95.7%) | 1.11 (0.96–1.29) | 1.13 (0.98–1.3) |
| Showcase | 12 (41.4%) | 91 (65.5%) | **1.58 (1.01–2.48)** | **1.66 (1.07–2.56)** |
| Subscription | 9 (31%) | 48 (34.5%) | 1.11 (0.62–2.01) | 1.18 (0.68–2.04) |
| External link | 21 (72.4%) | 101 (72.7%) | 1.01 (0.79–1.29) | 1.02 (0.79–1.32) |

Bold denotes statistical significance. PR: prevalence ratio; CI: confidence interval; a. adjusted for age, gender, year, quarter, and search query

**Supplementary Table 4.** Association between creator profession and health claim quality with unverified “doctors” categorized as physicians (N = 164)

| Health claim quality | Profession, n (%) |  | PR (95% CI) |  |
| --- | --- | --- | --- | --- |
|  | Medical Creators,<br>N = 33 | Non-Medical Creators,<br>N = 131 | Crude | Adjusted<sup>a</sup> |
| High | 16 (48.5%) | 5 (3.8%) | *Reference* | *Reference* |
| Poor or Moderate | 17 (51.5%) | 126 (96.2%) | **1.87 (1.34–2.6)** | **1.88 (1.37–2.58)** |

Bold denotes statistical significance. PR: prevalence ratio; CI: confidence interval; a. adjusted for age, gender, year, quarter, and search query

**Supplementary Table 5.** Associations between creator profession and financial conflicts with unverified “doctors” categorized as physicians (N = 174)

| Financial conflict | Profession, n (%) |  | PR (95% CI)<sup>a</sup> |  |
| --- | --- | --- | --- | --- |
|  | Medical Creators,<br>N = 35 | Non-Medical Creators,<br>N = 139 | Crude | Adjusted<sup>a</sup> |
| Any | 25 (86.2%) | 135 (93.1%) | **1.24 (1.03–1.49)** | **1.24 (1.05–1.48)** |
| Showcase | 12 (41.4%) | 91 (62.8%) | **1.91 (1.19–3.07)** | **1.99 (1.25–3.15)** |
| Subscription | 9 (31%) | 50 (34.5%) | 1.1 (0.64–1.89) | 1.26 (0.74–2.12) |
| External link | 21 (72.4%) | 101 (70.1%) | 1.22 (0.91–1.63) | 1.17 (0.88–1.55) |

Bold denotes statistical significance. PR: prevalence ratio; CI: confidence interval; a. adjusted for age, gender, year, quarter, and search query

